# T cell receptor sequencing specifies psoriasis as a systemic and atopic dermatitis as a skin-focused, allergen-driven disease

**DOI:** 10.1101/2021.07.14.21260435

**Authors:** Lennart M. Roesner, Ahmed K. Farag, Rebecca Pospich, Stephan Traidl, Thomas Werfel

**Author notes:** To whom correspondence should be addressed: Lennart Roesner, Hannover Medical School (MHH), Division of Immunodermatology and Allergy Research (OE6610), Carl-Neuberg-Str.1, 30625 Hannover, Germany. equal contribution (Lennart M. Roesner, Ahmed K. Farag).

## Abstract

**Background:** Atopic dermatitis (AD) and psoriasis represent two of the most common inflammatory skin diseases in developed countries. A hallmark of both diseases is T cell infiltration into the skin. However, it is still not clarified to what extent these infiltrating T cells are antigen-specific skin-homing T cells or unspecific heterogeneous bystander cells.

**Methods:** To elucidate this, T cells from lesional skin and from blood of 9 AD and 10 psoriasis patients were compared by receptor (TCR) sequencing. Therefore, peripheral blood mononuclear cells (PBMC) were cell-sorted according to expression of the cutaneous leukocyte antigen (CLA) into skin-homing (CLA^+^) and non-skin-homing (CLA^-^) subfractions. Aeroallergen-specific T cell lines were grown from AD patients’ PBMC in parallel.

**Results:** Intra-individual comparison of TCRB CDR3 regions revealed that clonally expanded T cells in skin lesions of both AD and psoriasis patients corresponded to skin-homing circulating T cells. However, in psoriasis patients, these T cell clones were also detectable to a larger extent among CLA^-^ circulating T cells. Up to 28% of infiltrating cells in AD skin were identified as allergen-specific by overlapping TCR sequences.

**Conclusions:** Our data shows that in line with the systemic nature of psoriasis, T cell clones that infiltrate psoriatic skin lesions do not exclusively possess skin-homing ability and are therefore most probably specific to antigens that are not exclusively expressed or located in the skin. T cells driving AD skin inflammation appear to home nearly exclusively to the skin and are, to a certain extent, specific to aeroallergens.

## Introduction

Atopic dermatitis (AD) and psoriasis are the most common inflammatory skin diseases with an incidence of 1-2% and 2-3% in adulthood, respectively ^1^. Regarding genetic risk factors, most loci identified are distinct for the two diseases while only a handful is shared ^2,3^. A hallmark of both diseases is T cell infiltration into the skin, but the phenotype and polarization status of infiltrating T cells differ substantially ^4^. The Th2-biased T cell response of AD and Th1/Th17-dominated cellular reactivity of psoriasis appear to be very stable; Eyerich and colleagues showed that in patients diagnosed with combined psoriasis and AD, the two diseases remain strictly within their opposing polarization profiles within the lesions ^5^. This argues for site-specific differences, probably regarding the skin composition and presence of specific antigens. AD is related to hypersensitivity reactions: Patients display a very heterogeneous pattern of allergic sensitization on a cellular and humoral level, leading to an individual pattern of antigens that may drive skin inflammation. In psoriasis, the existence of specific antigens is a matter of debate and the search is an ongoing task, with microbial and self-antigens having been proposed up to now ^6-10^.

To investigate the composition and distribution of the pathologically expanded T cells, T cell receptor (TCR) repertoire studies are the method of choice. T cell specificity is defined by the TCR CDR3 amino acid sequence which can be distinguished by its length (spectratyping) or the sequence itself. Before next-generation sequencing (NGS) became available ^11^, TCR repertoire analyzes were performed with technical limitations, but enabled first insights into the levels of AD ^12,13^ and psoriasis ^14-17^ T cell clonality.

Applying NGS to TCR sequencing was hampered for a long time by the genetically relatively long variable region and the large number of different V and J genes, which was solved by either multiplex-PCR ^18^ or RT-PCR ^19^ approaches. Both techniques bear drawbacks, namely primer bias or lack of proof-reading activity by the reverse transcriptase, respectively. Fortunately, the uncontrolled primer affinity bias of the multiplex-PCR was recently overcome ^20^.

As such, the first NGS TCR sequencing data on AD and psoriasis has become available, showing that general T cell clonality is comparable to healthy skin and much lower than is observed in cutaneous T cell lymphoma ^21-23^. Given the fact that the T cells present in healthy skin are also of a clonal origin since they originate from earlier inflammatory responses (so-called resident memory T cells (T_RM_) ^24-26^), this leads to the point that the T cell infiltrate in AD and psoriasis is, to a certain extent, oligoclonal. The question remains unanswered as to what extent the discovered T cells were truly licensed to enter the skin in order to encounter their respective antigen rather than being bystanders of the ongoing inflammation. T cells are actively recruited to the skin via homing molecules, first of all the cutaneous leukocyte antigen (CLA). Biochemically, CLA is a carbohydrate epitope of the sialic acid and fucose-modified platelet selectin glycoprotein ligand-1 (PSGL-1, a surface glycoprotein known to be expressed on the majority of peripheral blood leukocytes), induced by the fucosyl-transferase VII (FucT-VII). It functions a ligand for the selectins E, P, and L, thereby giving T cells access to inflamed cutaneous sites mediated by rolling on the superficial dermal epithelium of inflamed skin ^27,28^. Together with the lymphocyte function-associated antigen-1 (LFA-1) (which binds intercellular adhesion molecule-1 ICAM-1), very late antigen-4 (VLA-4) (binding the vascular cell adhesion protein-1 (VCAM-1), and chemokine ligands for chemokine, C-C motif, receptor (CCR) 10, CCR4, CCR6, and CCR8, a barcode system is formed that enables skin infiltration ^29^. It has been shown that in AD, CLA is expressed on all types of memory T cells, but not on naïve T cells. Further, CLA^+^ Th2 and Th22 cell frequencies correlate with the disease severity ^30^. The expression of Th2 cytokines includes IL-4 ^31^, IL-13 ^32^, IL-5 ^33^ IL-31, which links this T cells subset to the induction of itch ^34,35^. This type-2 inflammatory response is further fueled by thymic stromal lymphopoietin (TSLP), since CLA^+^ T cells express the respective receptor ^36^. In psoriasis, this subset is believed to establish the initial skin lesion and induce Th17/Th1/Th22 responses ^9,37^.

CLA^+^ T cells are considered a window to study cutaneous T cells without taking a biopsy, and serve therefore as a peripheral biomarker ^38^. E.g. Bakker et al. made use of CLA^+^ T cells as a peripheral biomarker when studying the effect of IL4R blockade in AD by dupilumab ^39^ and discovered that the biologic showed anti-inflammatory effects first and foremost on the CLA^+^ T cell fraction, underlining its pivotal role in AD ^40^.

In this proof-of-concept study, samples from blood and lesional skin were taken from AD patients and psoriasis patients. From the blood samples, several T cell subgroups were separated and analyzed separately, namely skin-homing T cells, non-skin-homing T cells, as well as allergen-specific T cells. Retrieving the sequences of TCR hypervariable domains of the T cells in each of these subgroups enabled us to re-identify respective blood-derived clones in the lesional skin in an intra-individual fashion. The T cellular skin infiltrate can therefore be described in a comprehensive manner.

## Results

### T cell repertoires of lesional skin of AD and psoriasis are mirrored predominantly by CLA^+^ blood T cells

Patients suffering from AD and psoriasis were recruited from the Dpt. of Dermatology and Allergy at Hannover Medical School, Hannover, Germany (Figure 1A,B). For TCR sequencing, genomic DNA was isolated from (a) a punch biopsy of lesional skin, (b) sorted peripheral blood skin-homing cells and (c) sorted peripheral blood non-skin-homing cells. Whilst non-skin-homing cells were defined as CD3^+^/CLA^-^, skin-homing T cells were identified by the fluorescent marker combination CD3^+^/CLA^+^ (Figure 1C). TCR sequencing of whole skin samples led to 4.98 million productive CDR3 sequences in total and 4.9-17.8 thousand unique sequences per sample (Figure 1D), revealing oligoclonal T cell expansion. Expanded clones were highly individual, as expected, with no common clonal variations among the different donors as shown by the CDR3 lengths (Figure 1E) and V/J-regions identified in the T cell subsets (Figure 1F and supplemental Figures 1,2,3,4,5).

**Figure 1.**
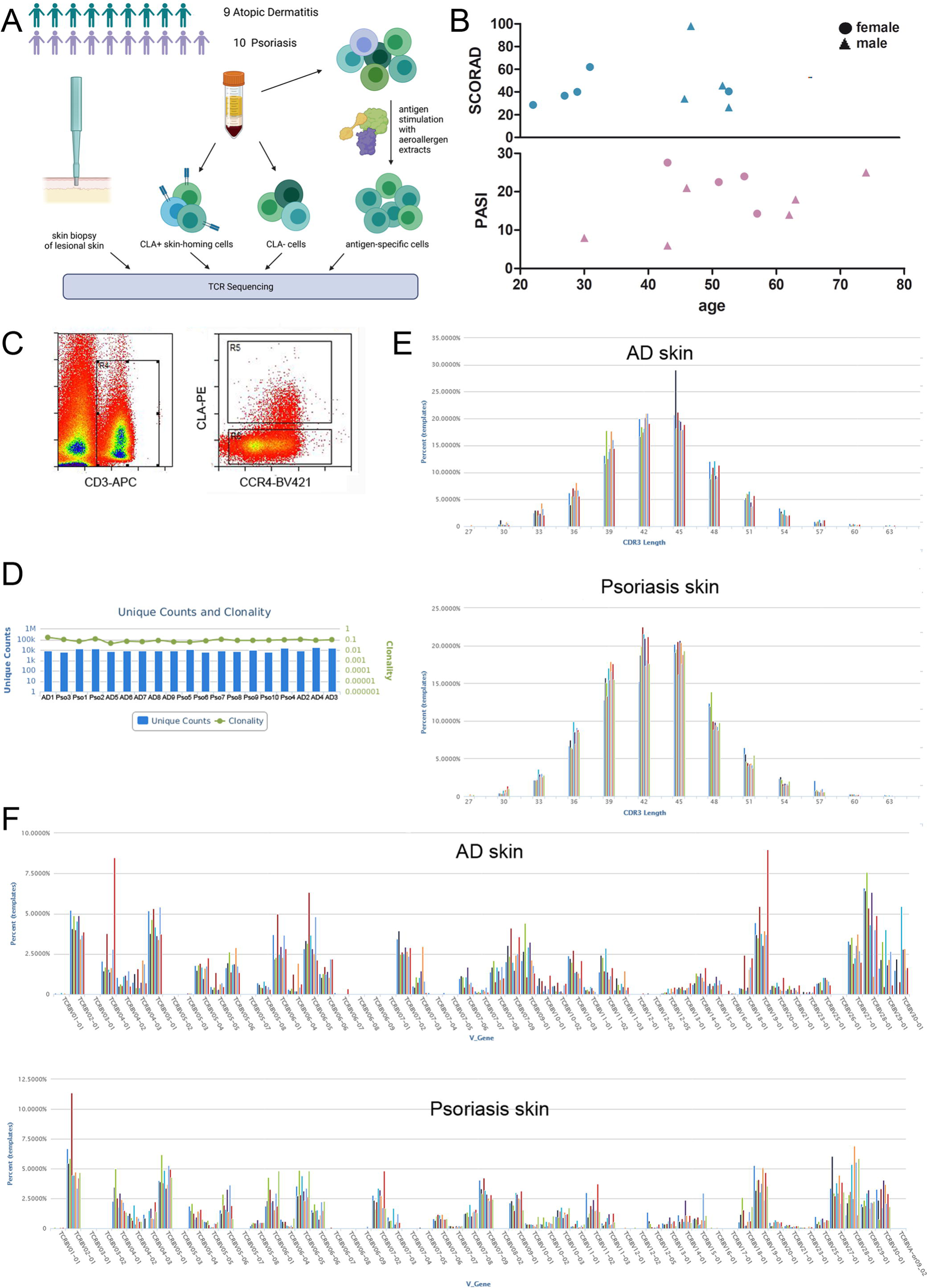
TCR repertoire analysis of lesional skin, skin-homing and non-skin-homing circulating T cells in patients suffering from AD and psoriasis. A, four samples per donor were analyzed by TCR sequencing: skin-infiltrating T cells, skin-homing T cells from PBMC sorted by CD3^+^/CLA^+^, non-skin-homing T cells from PBMC sorted by CD3^+^/CLA^-^, and T cell lines after antigen stimulation by allergen-extracts (only AD). B, patients’ characteristics including disease severity: SCORAD, Scoring Atopic Dermatitis; PASI, Psoriasis Area and Severity Index. C, Exemplary dot plots of the sorting strategy. D, Quality control TCR sequencing for skin-infiltrating T cells. E, CDR3 length distribution of skin-infiltrating T cells of AD and psoriasis skin samples. Coloured bars represent individual patients. F, V gene usage of skin-infiltrating T cells of AD and psoriasis skin samples. Respective data for subsets of circulating T cells depicted in the supplementary material.

The skin-homing ability of T cells is mediated by CLA, therefore, T cells in lesional skin are also expected to be present in the CLA^+^ fraction of circulating cells. The intra-individual overlap of T cell repertoires across lesional skin and circulating CLA^+^ and CLA^-^ subpopulations are shown for AD and psoriasis (Figure 2A, B). In both AD and psoriasis, lesional skin shared more clones with the skin-homing (CLA^+^) fraction than with the CLA^-^ fraction, without detectable inter-individual clonal bias in terms of their V/J gene usage (supplemental Table 1). This suggests that the majority of skin-infiltrating cells target antigens that are encountered in or on the skin. Nevertheless, the overlap of skin-infiltrating T cell clones with non-skin-homing CLA^-^ T cells in the circulation was higher in psoriasis compared to AD (Figure 2C).

**Figure 2.**
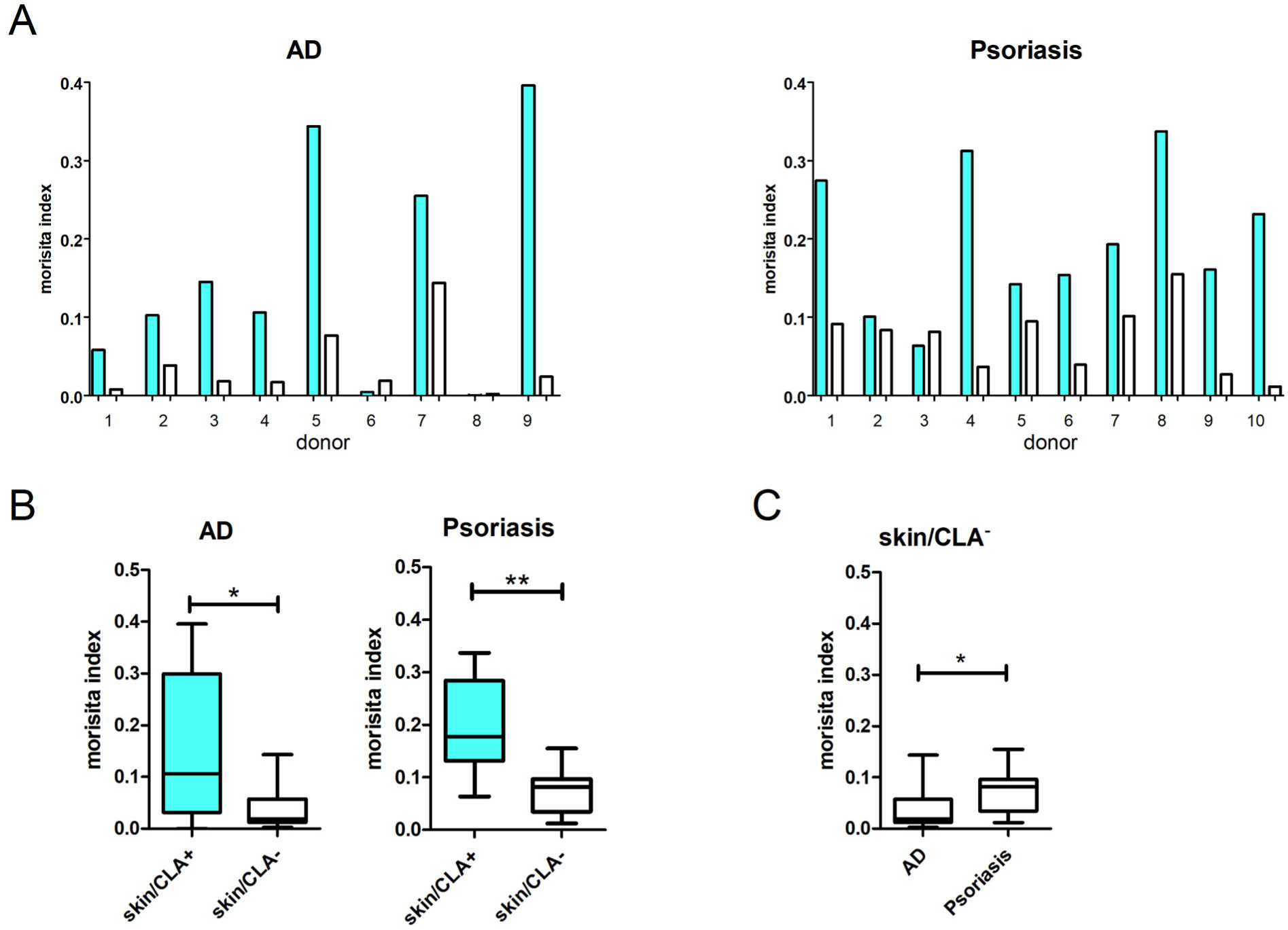
Comparative TCR repertoire analysis of lesional skin T cells with skin-homing and non-skin-homing circulating T cells. The TCR repertoire of lesional skin was compared with the skin-homing (CLA^+^, grey) and the non-skin-homing (CLA^-^, white) fraction of PBMC intra-individually. The overlap of the respective repertoires is displayed by the morisita index. A, bar charts represent single donors’ overlap between lesional skin T cells with either CLA^+^ or CLA^-^ circulating T cells. B, Overlap of lesional skin T cells with either CLA^+^ or CLA^-^ circulating T cells. \*\**P*<.01, Wilcoxon matched pair test. C, Overlap of lesional skin T cells with CLA^+^ or CLA^-^ circulating T cells in AD compared to psoriasis. *P*=.072, Mann-Whitney test. Whiskers of Box plots represent min / max values. AD n=9, Psoriasis n=10.

### Highly clonally propagated T cells in AD lesional skin are largely derived from CLA^+^ blood T cells, in contrast to psoriasis

In the following we focused on T cells that were clonally propagated, representing *bona fide* disease-driving T cells. In Figure 3A,B the frequencies of each donors’10 most frequent clones detected in skin, CLA+ and CLA-fractions are shown. Interestingly, we detected comparable frequencies of clonally expanded T cells in the samples derived from AD and psoriasis skin (Figure 3A,B, up to 4.8% in AD and 6.5% in psoriasis). The two most frequent clones in each patient’s skin sample were present above 1% in median, arguing for a strong T cell response to certain specific antigens (Figure 3A). Comparing the skin with the CLA^+^ and CLA^-^ PBMC fraction, T cell clones with the highest frequency were detected in the CLA^+^ T cell fraction in AD, with up to 7% (median 1.3±2.5 SD), followed by the skin (Figure 3A). This supports the widespread opinion that antigens recognized via the skin are of high importance in AD. In psoriasis, T cell clones with the highest frequency were detected in the CLA^-^ T cell fraction of each patient, with up to 8% (median 3.3±2.7 SD), followed by the skin (Figure 3A). The clonality of the 10 most frequent T cell clones of the CLA^-^ fraction was thereby significantly higher in psoriasis compared to AD, while vice versa the clonal expansion of the CLA^+^ fraction was higher in AD compared to psoriasis (Figure 3B, supplemental Figure 6). Adding to this, T cell clones that are shared between lesional skin and the CLA^-^ fraction do not occur in high frequency in AD. In psoriasis, these are often detected in frequencies above 0.5% (supplemental Figure 7).

**Figure 3.**
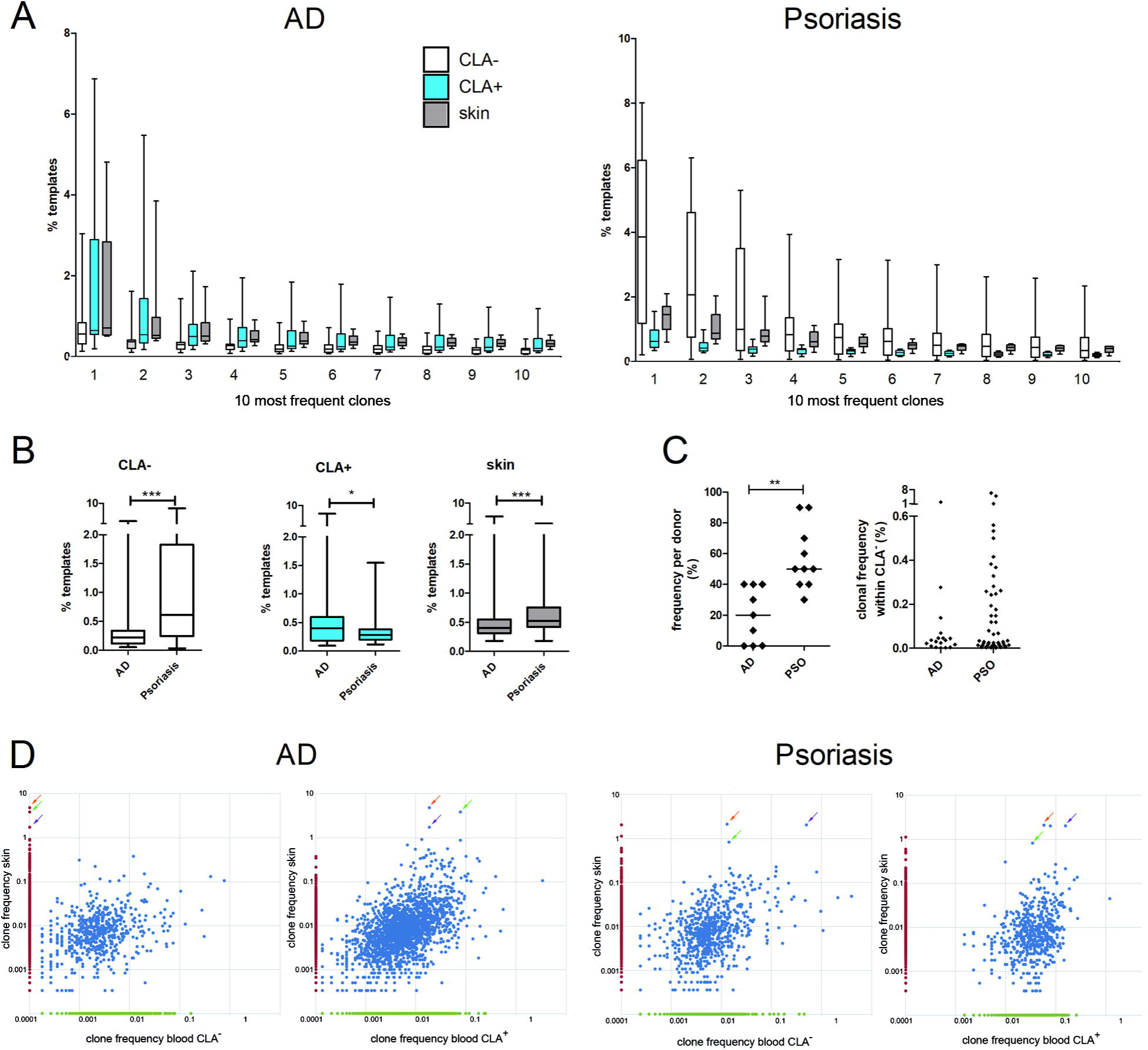
TCR repertoire analysis of highly clonally propagated T cells. A, Clonal frequencies of the donors’ 10 most frequent T cell clones in the non-skin-homing (CLA^-^, white) the skin-homing (CLA^+^, light grey) fraction of PBMC, or skin-infiltrating T cells (dark grey). The 10 most frequent clones per donor are denoted as 1-10 on the x-axis and the respective frequency on the y-axis. B, Combined clonal frequencies of the donors’ 10 most frequent T cell clones. C, left: Percentage of T cell clones is depicted, that are shared between lesional skin and the CLA^+^ T cell fraction and in addition present in the CLA^-^ T cell fraction (focusing each donors’ 10 most frequent T cell clones). Right: respective clone frequencies. D, exemplary dot plots of donors AD1 and PSO2 demonstrating that highly clonally propagated skin T cells (arrows) in AD derive from CLA^+^ circulating T cells, while in psoriasis these are also found in the CLA^-^ circulating T cell fraction. Mann-Whitney test. Whiskers of Box plots represent min / max values. AD n=9, Psoriasis n=10.

This may indicate that driver T cells in psoriasis recognize antigens that are not exclusively encountered at the skin, but also at other body sites. The multiple co-morbidities of psoriasis, such as arthritis, would support this theory.

Our findings on the 10 most frequent clonally expanded T cell clones of lesional skin that are also present in the CLA^+^ fraction further support this (Figure 3C): In AD, only a few of these T cell clones were detected also in the CLA-fraction (15% in median, with low clonal frequencies). In psoriasis, these were detectable to 50% in median also in the CLA-fraction. On the one hand, this underlines the assumption, that antigens which are recognized first and foremost via the skin are of high importance in AD. On the other hand this may indicate that in psoriasis bona fide multi-organ-homing T cells play an important role. Such multi-organ homing T cell clones occurred in high frequencies above 0.1% in our set of patients nearly exclusively in psoriasis compared to AD (supplemental Figure 8).

To further visualize this opposing picture presented by the two inflammatory skin diseases, two examples are shown in Figure 3D. The most common skin T cell clone in AD was found in patient 1 with a frequency of 4.8% (orange arrow). While also being frequently detectable in the CLA^+^ blood T cell fraction (0.019%), it was absent in CLA^-^ blood T cell fraction. On the other hand, the most common skin T cell clone in psoriasis patient 2 was found with a frequency of 2.1% and was also present in the CLA^+^ and CLA^-^ sample with frequencies of 0.06% and 0.01% respectively.

### Aeroallergen-specific T cells are part of the AD skin infiltrate

To investigate if (and to what extent) aeroallergen-specific T cells are part of the skin infiltrate, allergen-specific T cell lines were generated from PBMC of 7 out of the 9 AD donors of this study. The respective allergen extracts applied for *in vitro* stimulation were chosen according to each patient’s IgE sensitizations (Supplemental Table 2) comprising of house dust mite (HDM), grass pollen mix, birch pollen, and rye pollen as a proof-of-concept. T cell line specificities were confirmed by re-stimulation proliferation testing. If a T cell clone, defined by its CDR3, was present and propagated in the allergen-reactive T cell line with at least a 3-fold higher frequency compared to unstimulated PBMC of the same donor, or even absent in the latter, it was regarded as allergen-specific.

These aeroallergen-specific T cells were compared on an intra-individual level to skin-infiltrating T cells. Focusing on clonally propagated T cells in lesional skin (with frequencies > 0.1% in skin), 51 different aeroallergen-specific CDR3 sequences were re-identified from the T cell lines (Supplemental Table 3). While there was strong variation between individuals, these aeroallergen-specific T cells made up to 28.3% of clonally propagated T cells in lesional skin (Figure 4). Interestingly, these CDR3 were further on detectable among circulating blood cells, but in significantly higher frequencies in CLA+ compared to CLA-samples (median±SEM: CLA^+^ 1.0±0.5%; CLA^-^ 0.0±1.3%; p=.0001).

**Figure 4.**
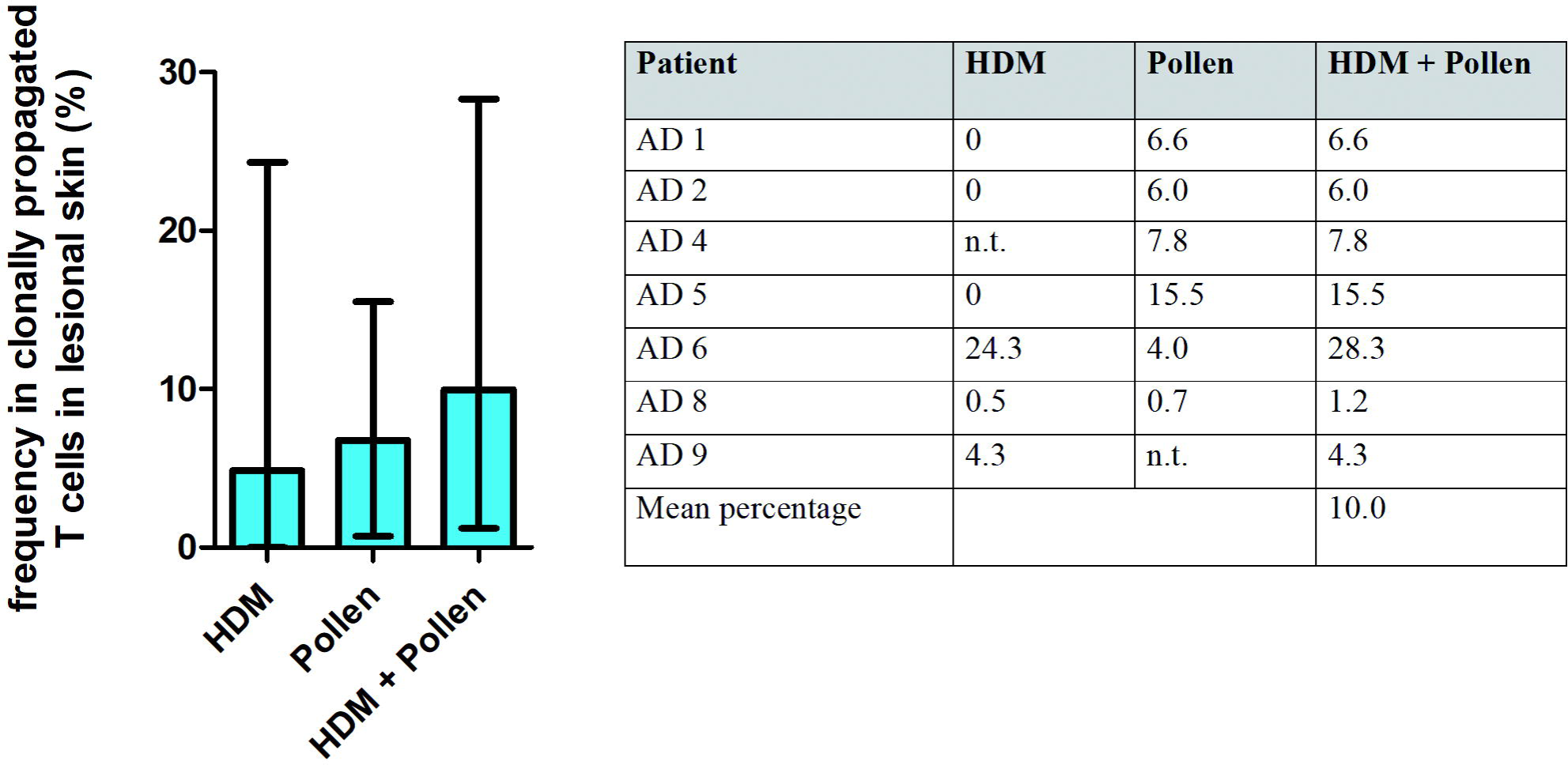
Percentage of allergen-specific clones within clonally expanded T cell clones in lesional AD skin. Only skin T cell clones with frequencies above 0.1% were investigated. T cell clones were defined as allergen-specific on the basis of aeroallergen-reactive T cell lines generated from each patient’s blood. If a T cell clone, defined by its CDR3, was present and propagated in the allergen-reactive T cell line with at least a 3-fold higher frequency compared to unstimulated PBMC of the same donor, or even absent in the latter, it was regarded as allergen-specific. Data is presented as mean with range (left), and as individual data in the table (right). HDM, T cell clones with specificity for house dust mite extract. Pollen, T cell clones with specificity for birch, grass, or rye pollen extract. Standard deviations: HDM: 9.7, pollen: 4.9, HDM+pollen: 9.2.

Some clonal T cells were found to proliferate in response to several different pollen antigens (grass, birch, rye) in the respective T cell line, suggesting allergen-cross-reactive T cell clones (Supplemental Table 3).

T cell clones present in the skin with frequencies below 0.1% were considered as putatively less disease-driving, possibly bystander cells, and therefore excluded from this analysis. This proof-of-concept approach shows that a distinct subset of T cells infiltrating lesional AD skin can be identified as aero-allergen-specific.

## Discussion

When applying TCR sequencing to skin, caution should be taken in comparing the clonality of T cells within different sample entities. Healthy skin does indeed harbor T cells; so-called resident memory T cells (T_RM_) ^24^. These are believed to originate from former pathogen attack and subsequent inflammatory response, building a local memory that is able to respond quickly in case of a second pathogen encounter ^25,26^. Therefore it is obvious that healthy skin harbors a set of oligoclonal T cells (with specificity to a limited set of skin-pathogen antigens) and not the broad range of theoretically 10^14^-10^17^ different TCRs found in the bloodstream. Recently, TCR deep sequencing technology has been applied to investigate the generation of healthy skin T_RM_. It could be demonstrated that there is a common clonal origin of central (T_CM_) and T_RM_ following skin immunization, demonstrating that the skin has the inflammatory potential to act as a route of immunization ^23,25^. In inflamed skin, whilst T_RM_ may be outnumbered by infiltrating T cells, their clonality may be higher, lower, or in the same range as the T_RM_, depending on the number and dominance of different disease-specific antigens. Therefore, a comparison of clonality levels to healthy skin may sometimes be misleading. To shed light on those T_RM_, that are generated as residuals from episodes of chronic inflammatory skin diseases like AD and psoriasis, it is still an unmet need to contrast these to infiltrating T cells of acute lesional skin.

Prinz and colleagues have previously suggested that psoriasis vulgaris has a common antigen since T cells with conserved TCRB CDR3 sequences were found in identical twins by Sanger sequencing of T cell clones ^41^. Moreover, TCR analysis of psoriatic lesional skin in patients with different psoriasis associated co-morbidities has been performed ^15-17^, detecting shared clones between different affected body sites ^17^. However, in these approaches the technology used was limited regarding the visualization of the billions of clones forming the TCR repertoire.

Another study performing TCR deep sequencing in skin of patients with cutaneous T cell lymphoma (CTCL) ^42^ demonstrated that this technique could accurately diagnose CTCL in all disease stages, and further identified mature T cells as the cell of origin. Among their results, they showed that the clonality levels in AD and psoriatic skin lesions are far less than in CTCL. In general, our measurements confirm this finding, but when taking a deeper look and taking into account the different skin-related blood compartments, we were able to detect oligoclonality in a subset of T cells. Highly propagated T cell clones were found in frequencies up to 4.8 % in AD patient skin and were also detectable as CLA-expressing cells in the bloodstream. It can be assumed that these putatively disease-driving clones are therefore licensed to enter the skin, most likely after encountering their cognate skin-borne antigen in the lymphoid organs.

In the psoriasis patients analyzed, the most abundant clones in skin corresponded to the CLA^-^ blood fraction, suggesting that these cells were not exclusively skin-specific, and may target other body sites. Even more compellingly, circulating cells that did not express the skin homing factor CLA had higher clonality levels. This matches to the observation made by Matos et al., reporting that active psoriatic sites harboured more polyclonal T cells compared to clinically resolved skin ^22^. Interestingly, putative T cell antigens in psoriasis have been reported to be produced by non-skin-cells, which could explain the lack of CLA ^43,44^.

A possible factor that might influence the results reported here is possible recirculation of T cells from the skin back into the blood. This concept of recirculation has been postulated several times ^45-47^ based on observations from clinical trials with LFA-1-antibodies to block skin-infiltration ^48,49^. These studies observed accumulating and bona fide recirculating T cells in the blood, which were CLA^+^. In this case, the circulating CLA^+^ fraction investigated here could be considered as a prospective and retrospective peripheral cellular biomarker, mirroring cutaneous T cells ^38^.

Comparing T cells of lesional skin to circulating T cells sorted by their CLA expression has revealed clear differences between the two model diseases of psoriasis and AD. The T cell repertoire of lesional AD skin shows a high overlap with the CLA^+^ and allergen-specific circulating T cells, and frequent T cell clones were found exclusively within these subsets. This underlines the dominant inflammatory role of skin-homing T cells in AD. Indeed we have recently demonstrated that exposure to allergens via the surrounding air leads to strong eczema flaring in allergen-exposed skin, highlighting the direct inflammatory route that exists via the skin ^50^.

Revealing that clonally expanded T cells found in psoriatic skin are not exclusively skin-homing may reflect a systemic inflammation; it may be hypothesized that expanded T cell clones common to both skin and the CLA^-^ circulating fraction represent memory T cells specific to an epitope present in both the skin and another anatomical location. For example cross-reactive autoantibodies directed against antigens found in skin and joints in psoriatic arthritis have been described ^51^. Psoriasis is associated with a risk of developing psoriatic arthritis ^52^ and other inflammatory disorders and is therefore considered a more systemic disease. The distribution of clonally expanded T cells reported here supports this idea and provides new information about the composition of the heterogeneous population of T cells. Although CLA^+^ T cells were also proven to play a role in the initiation of psoriatic skin lesions ^37^, our results suggest that CLA is not a robust marker for clonally expanded T cells in this disease.

It is surprising that the skin infiltrate of AD patients is dominated by highly frequent clonally expanded T cells (some T cell clones make up to 4.8% of the AD skin infiltrate). AD patients are known to be sensitized against numerous allergens, each harboring several epitopes, which would lead to a heterogeneous T cell population. In order to assign clonally expanded T cells to antigen-specificities, TCL were generated. One technical drawback of this approach is that by starting with a limited number of cells (1*10^6^) rare T cell clones may be overlooked. Furthermore, differentiated T cells that are believed to be strong cytokine producers have been reported to bear less proliferative capacity and therefore may be overwhelmed by others during TCL generation ^53^. Completely exhausted cells may also be lost during culture due to activation-induced cell death, thereby skewing the T cell line and leading to artificial clonal frequencies. Therefore it can be assumed that our measurement of allergen-specific skin-infiltrating T cells is an underestimation. Taken together, this approach shows that, in AD, clonally expanded T cell clones which react to allergens mirrored by the individual patients’ IgE sensitization profiles resemble a distinct proportion of the inflammatory skin infiltrate. Further T cell targets are most probably microbial antigens ^54-56^ and autoantigens ^57^. Nevertheless, the technical approach of TCR sequencing circumvents the inherent biases of cultivating skin-derived T cells. T cell responses towards putative antigens with relevance in psoriasis ^6-8^ could be estimated this way, too.

Frequent clones may serve as a promising marker clone in single patients on an individual level. By following patient-specific marker T cell clones during the course of the pollen season or an allergen-specific immunotherapy, a deeper understanding of the individual response to allergen and treatment could be achieved. We believe that T cell repertoire based biomarkers, if developed, can be a powerful tool in the field of precision diagnostics compare ^58^. Adding to that, knowledge of allergen-specific clones in AD can create new targets for a more precise, causative treatment.

## Materials and Methods

### Patients

Whole blood was taken from adult patients suffering from AD or psoriasis alongside 4 mm punch biopsies from full-thickness lesional inflamed skin. AD patients were defined following the criteria by Hanifin and Rajka ^59^. No patient in this study was under any local or systemic treatment when the samples were taken. This study was performed according to the Declaration of Helsinki, approved by the Ethics Committee of Hannover Medical School (No. 3362), and all patients gave their written informed consent. Patients showed moderate to severe disease activity; patient data and the appropriate severity scores (SCORAD/PASI) are depicted in Figure 1.

### Nucleic acid isolation

Skin samples were homogenized using the TissueRuptor (Qiagen, Valencia, CA, USA). Genomic DNA was isolated from skin extracts and sorted blood cells using the NucleoSpin Tissue Mini Kit (Macherey & Nagel, Dueren, Germany) according to manufacturer’s instructions. DNA integrity was confirmed by Bioanalyzer (Agilent, Santa Clara, CA, USA).

### Cell sorting

PBMC were isolated by Ficoll density centrifugation from all blood samples. Cells were stained with anti-CD3-allophycocyanin antibodies (BeckmanCoulter, Brea, CA, USA), anti-chemokine (C-C-motif) receptor 4 (CD194, CCR4)-BrilliantViolet421 antibodies (Biolegend, San Diego, CA, USA) and anti-CLA antibodies coupled to phycoerythrin (Miltenyi Biotech, Bergisch Gladbach, Germany). Cell sorting into CD3^+^/CLA^+^ and CD3^+^/CLA^-^ fractions was performed using either the MoFlo XDP (Beckman-Coulter, Fullerton, CA, USA) or FACSAria Fusion (BD Biosciences, San Jose,CA,USA) in the cell sorting core facility of Hannover Medical School.

### Identification and quantification of TCR sequences of antigen specific cells

In order to identify antigen specific clones, specific T cell lines (TCL) were grown for three weeks from PBMC extracted from the same set of patients in the presence of allergen extracts (purchased at ALK, Horsholm, Denmark, Citeq Biologicals, Groningen, The Netherlands, and the National Institute for Biological Standards and Control (NIBSC), South Mimms, Great Britain) according to an established protocol ^60^. Allergen extracts were selected based on each patient’s specific IgE titers as measured by CAP Feia (Thermo Scientific, Massachusetts, USA) towards the tested allergens, namely birch pollen, house dust mite, rye pollen, and grass pollen. Allergen-specific IgE-titers of >3.19 kU/I were regarded as relevant. Antigen-specificity of each cell line was approved after 21 days by re-stimulation testing. Therefore, respective antigen-presentation by irradiated autologous antigen-presenting cells was followed by proliferation testing by ^3^H-thymidine incorporation. Finally, DNA was extracted and used for TCR sequencing. PBMC cultured in the absence of antigens served as controls.

### Next generation TCR sequencing

Sequencing was performed using the Adaptive Biotechnologies (Seattle, WA, USA) ImmunoSEQ Human T-cell Receptor Beta (hsTCRB) kit and service. This technique is based on DNA templates and applies a tightly controlled multiplex PCR followed by NGS on Illumina MiSeq and NextSeq (Illumina, San Diego, CA) analyzers ^20,61^. NGS TCR sequencing was performed with 100,000 reads (survey resolution) including V, D, and J family sequencing for each CDR3,filtering out non-productive sequences. All samples passed quality control at Adaptive Biotechnologies.

### Data analysis

The bioinformatics pipeline of the ImmunoSEQ platform for TCRB CDR3 analysis was applied, using in-silico methods to eliminate potential amplification bias. Data were analyzed using the ImmunoSEQ analyzer 2.0 and 3.0 (Adaptive Biotechnologies) to display the overlap of the respective repertoires by morisita index. This index describes the overlap between two samples by taking into account both the number of clonotypes as well as their frequencies within the repertoire. An index of 1 represents complete overlap or identical repertoire and an index close to 0 represents two very different repertoires. The Morisita index is considered a robust method especially for TCR repertoire studies (reviewed in ^62^).

### Statistics

Statistical analyses were performed using Graphpad Prism 5 (Graphpad, San Diego, CA, USA). Differences were considered significant if *P*<.05.

## Data Availability

All data are available in the main text or the supplementary materials.

## Acknowledgments

We thank Mrs. Petra Kienlin and Mrs. Gabriele Begemann for excellent technical assistance. We thank Dr. Lutz Wiehlmann, Dr. Colin Davenport and Mrs. Marie Dorda from the Research Core Unit Genomics (RCUG), Hannover Medical School, Germany, for fruitful discussion on TCR sequencing, quality control and perfect sample handling. Further we thank Dr. Matthias Ballmaier and his team at the core facility for cell sorting, Hannover Medical School, Germany. Graphical Abstract was created with Biorender. We thank Tom Macleod, University of Leeds, for English proofreading.

## Notes

### Competing Interest Statement

LMR declares grants to his institution and personal fees from Novartis. AKF is an employee of Boehringer Ingelheim GmbH. ST received consultancy fees from LEO Pharma, Lilly, and La Roche Posay. RP declares that she has no competing interests. TW has received institutional research grants from LEO Pharma and Novartis, has performed consultancies for Abbvie, Janssen, Galderma, LEO, Sanofi-Genzyme, and Novartis, he has also lectured at educational events sponsored by Abbvie, Janssen, Celgene, Galderma, LEO Pharma, Sanofi and Novartis and is involved in performing clinical trials with various pharmaceutical industries that manufacture drugs used for the treatment of and atopic dermatitis.

### Funding Statement

Institutional funding.

### Author Declarations

The study was approved by the Internal Review Board of Hannover Medical School.

