## Supplemental Figure for "T cell receptor sequencing specifies psoriasis as a systemic and atopic dermatitis as a skin-focused, allergen-driven disease"

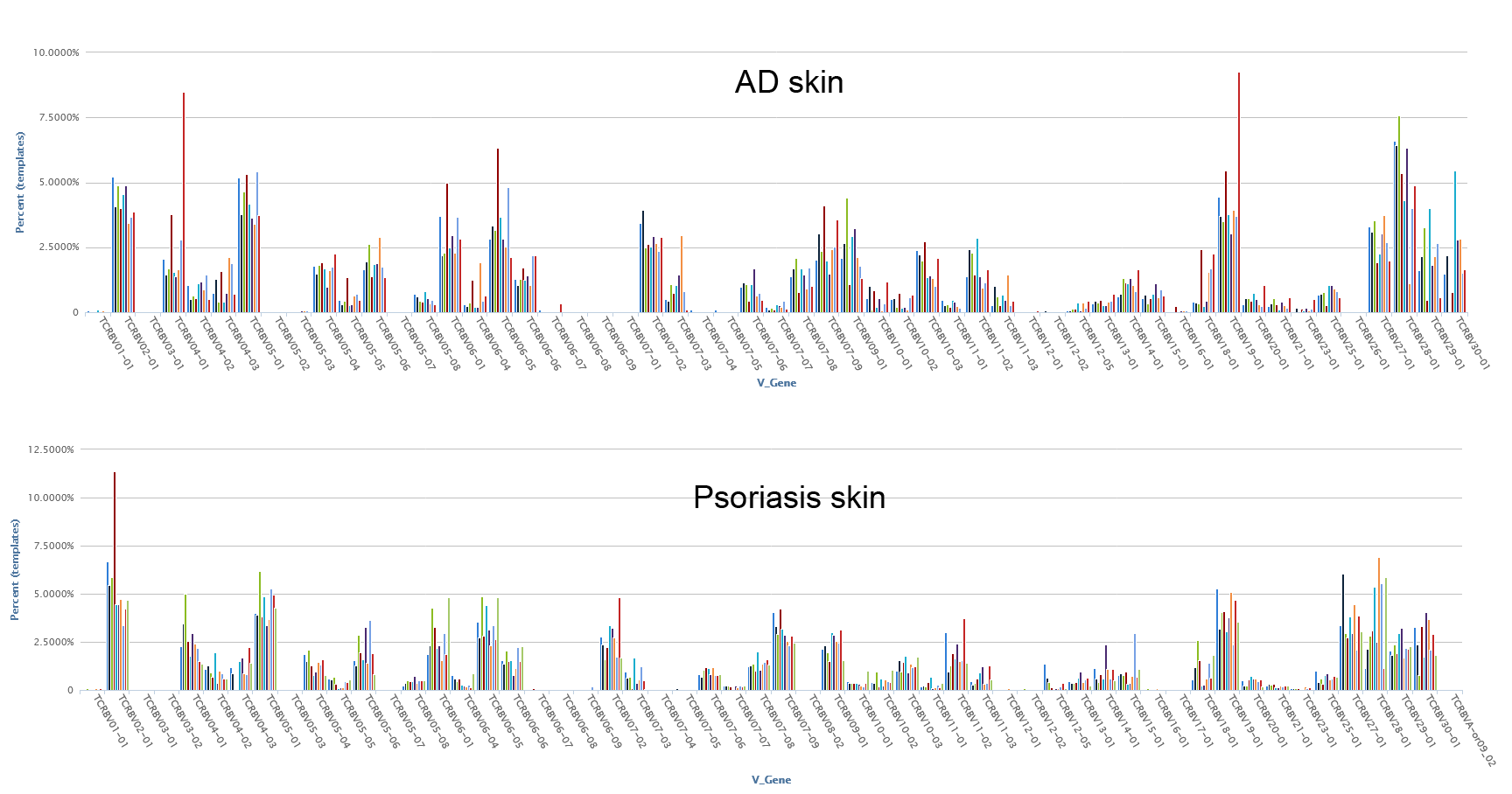

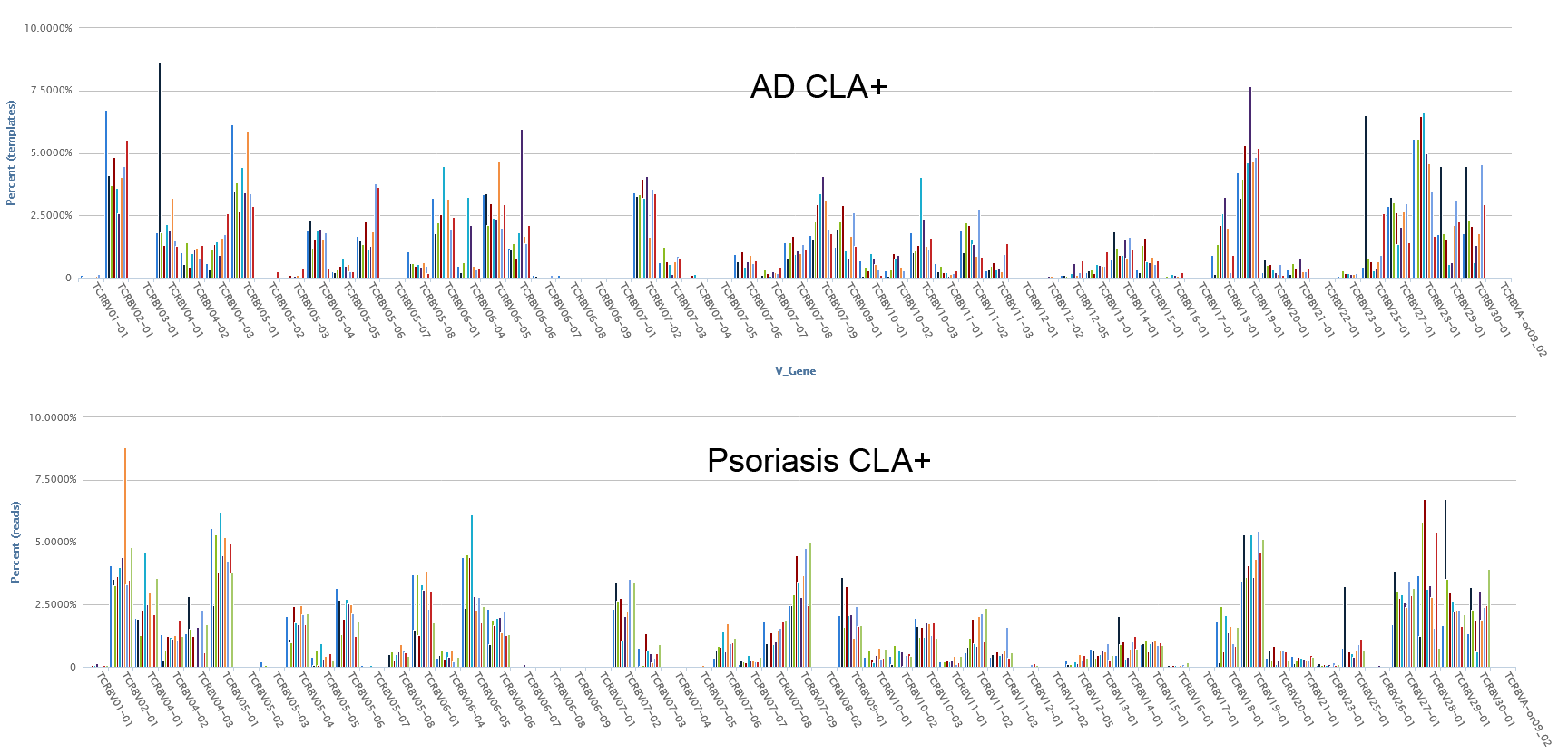

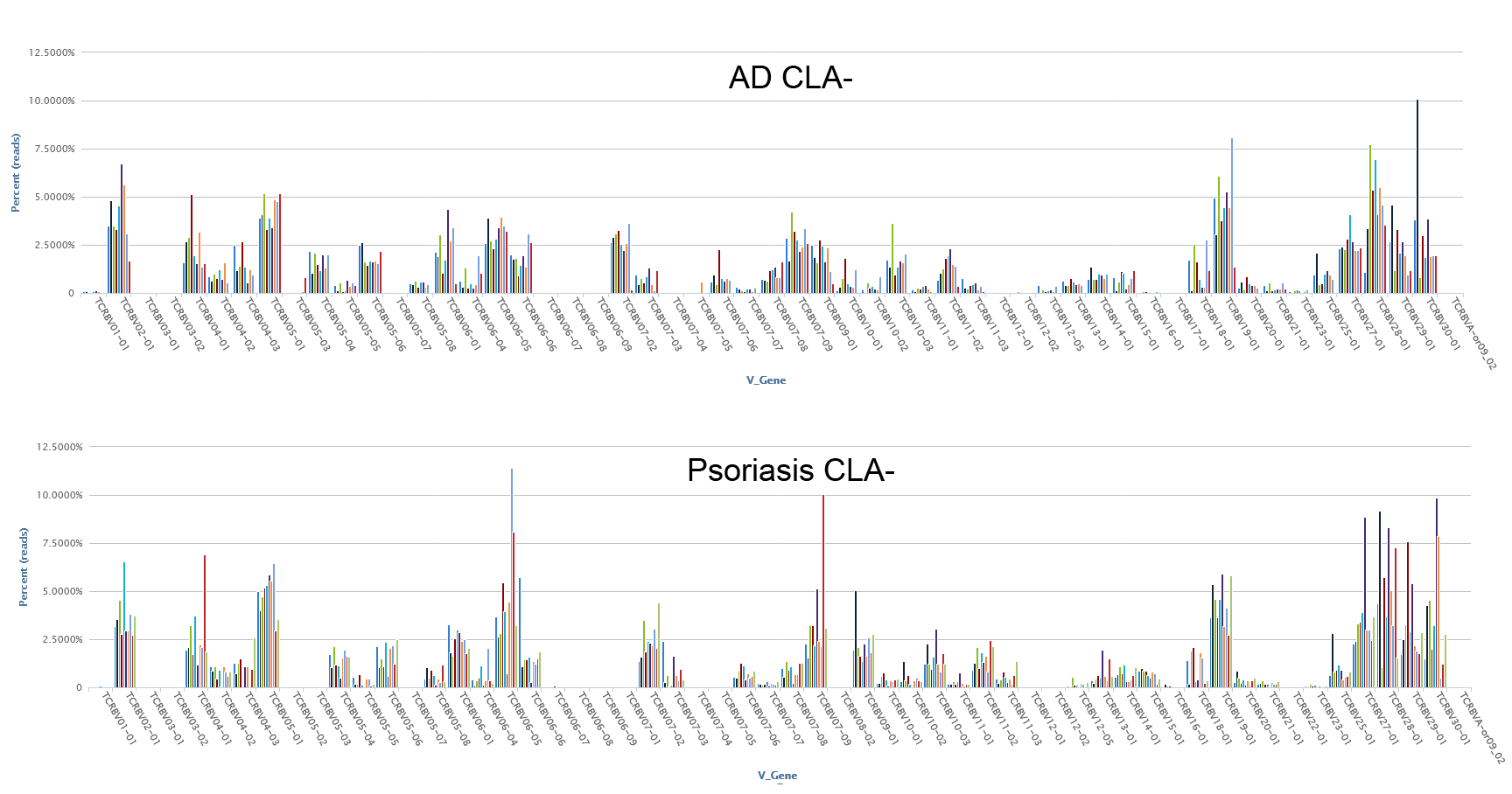
 **Supplemental Figure 1.** TRBV gene usage among skin-infiltrating (skin), skin-homing (CLA^+^) and non-skin-homing (CLA^-^) T cells. Coloured bars represent individual patients.

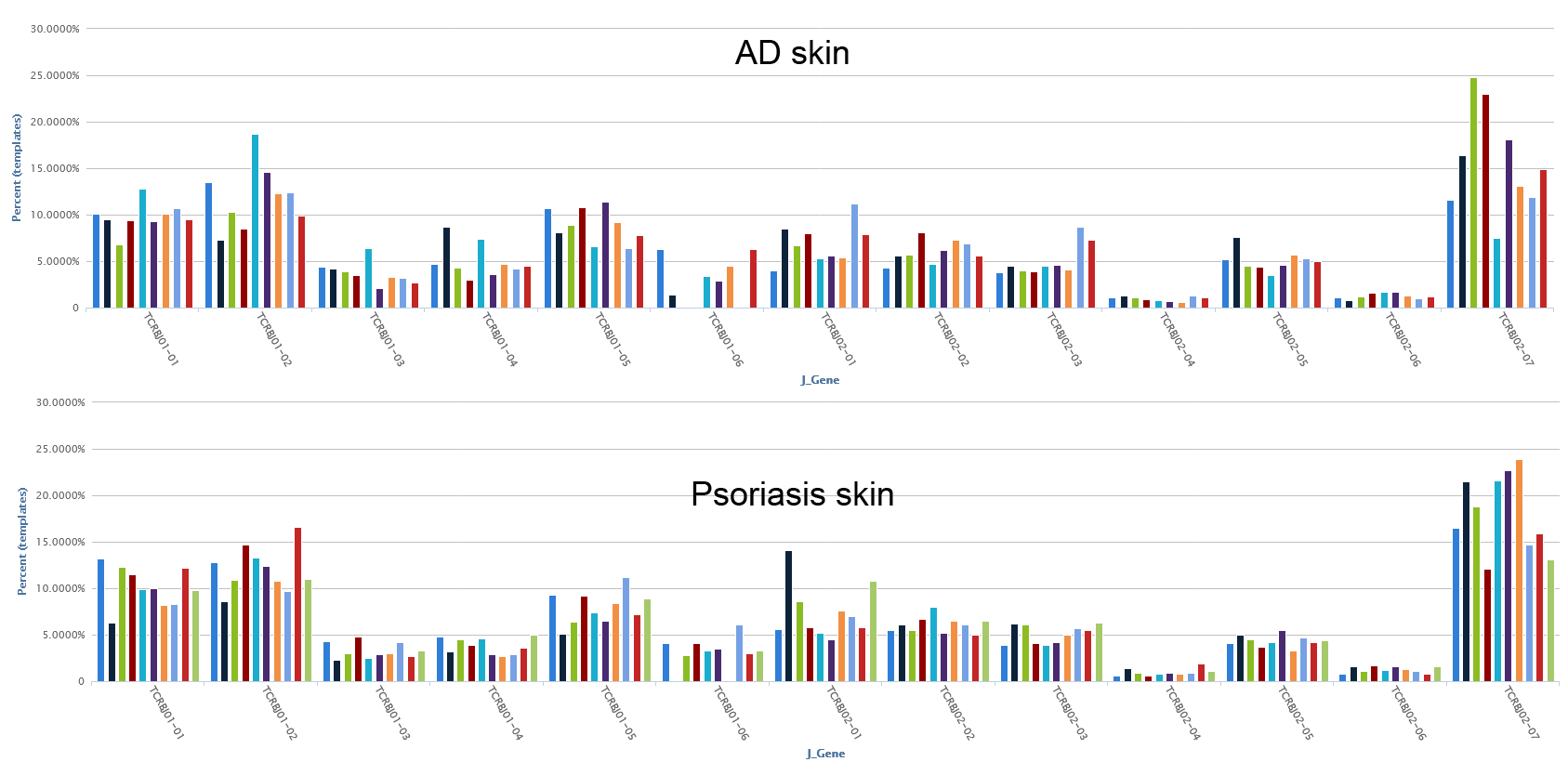

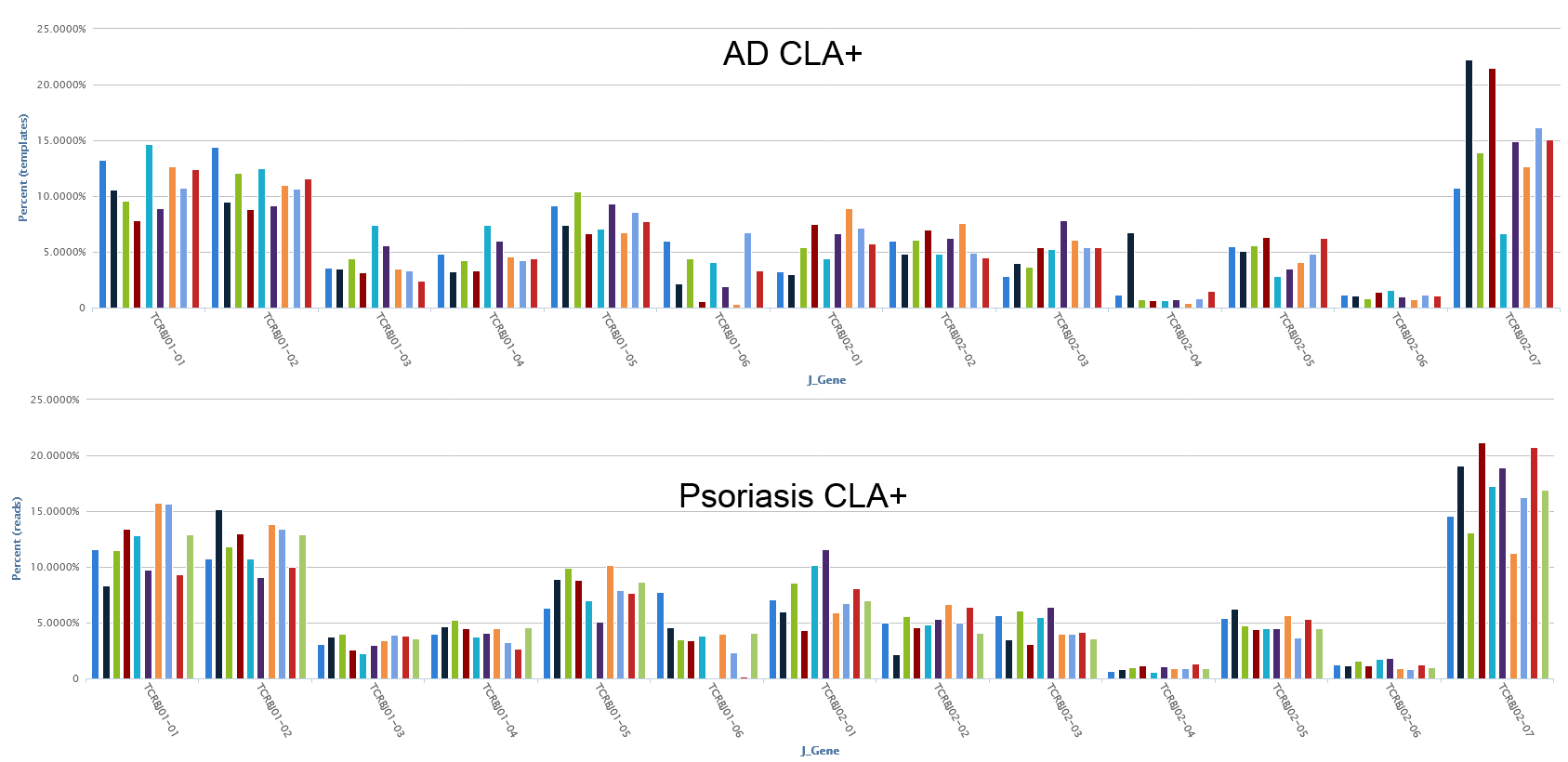

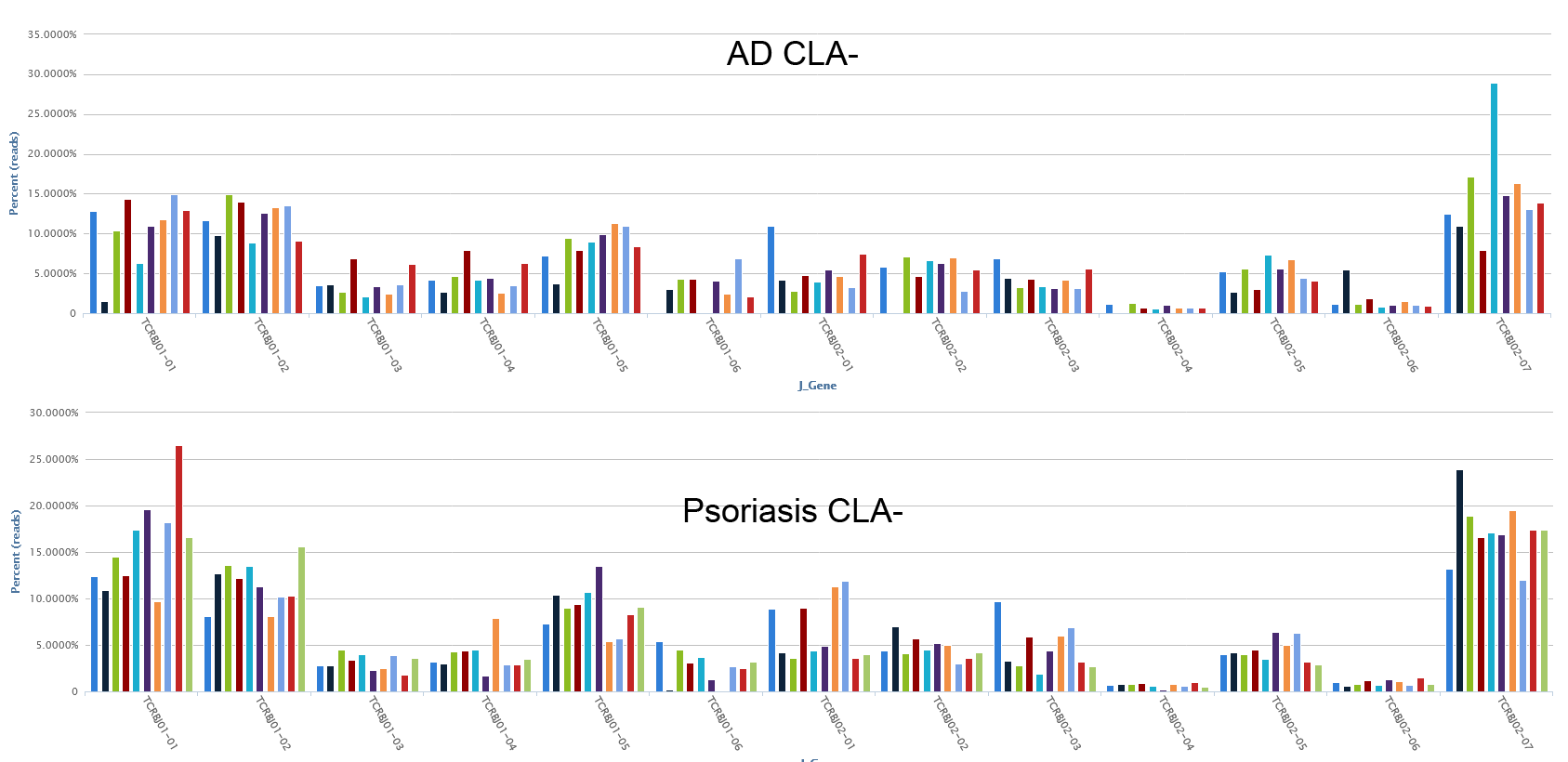
 **Supplemental Figure 2.** TRBJ gene usage among skin-infiltrating (skin), skin-homing (CLA^+^) and non-skin-homing (CLA^-^) T cells. Coloured bars represent individual patients.

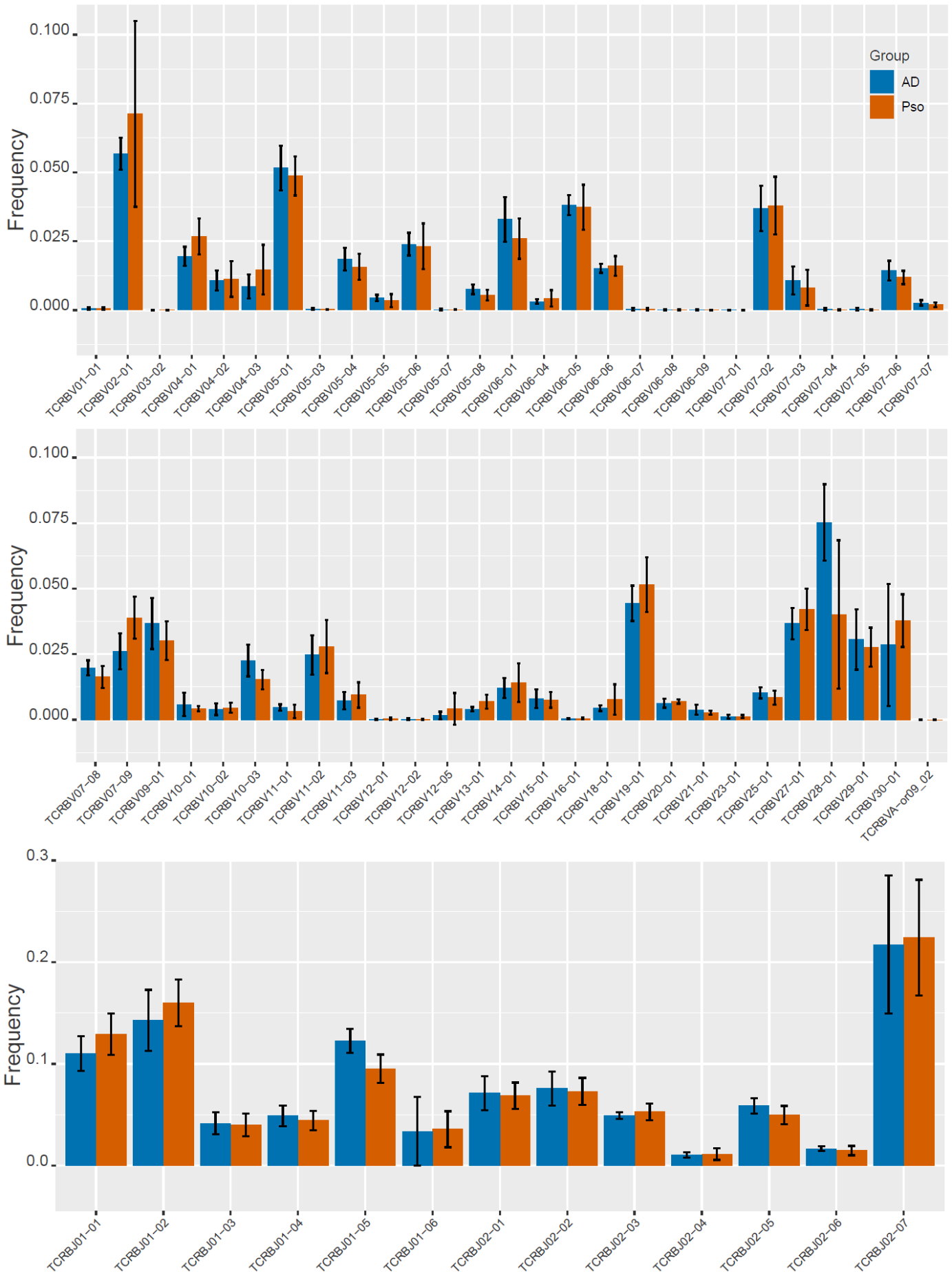

**Supplemental Figure 3**: Percentages of the TRB genes used by skin-infiltrating T cells in AD compared to psoriasis. AD n=9, Psoriasis n=10.

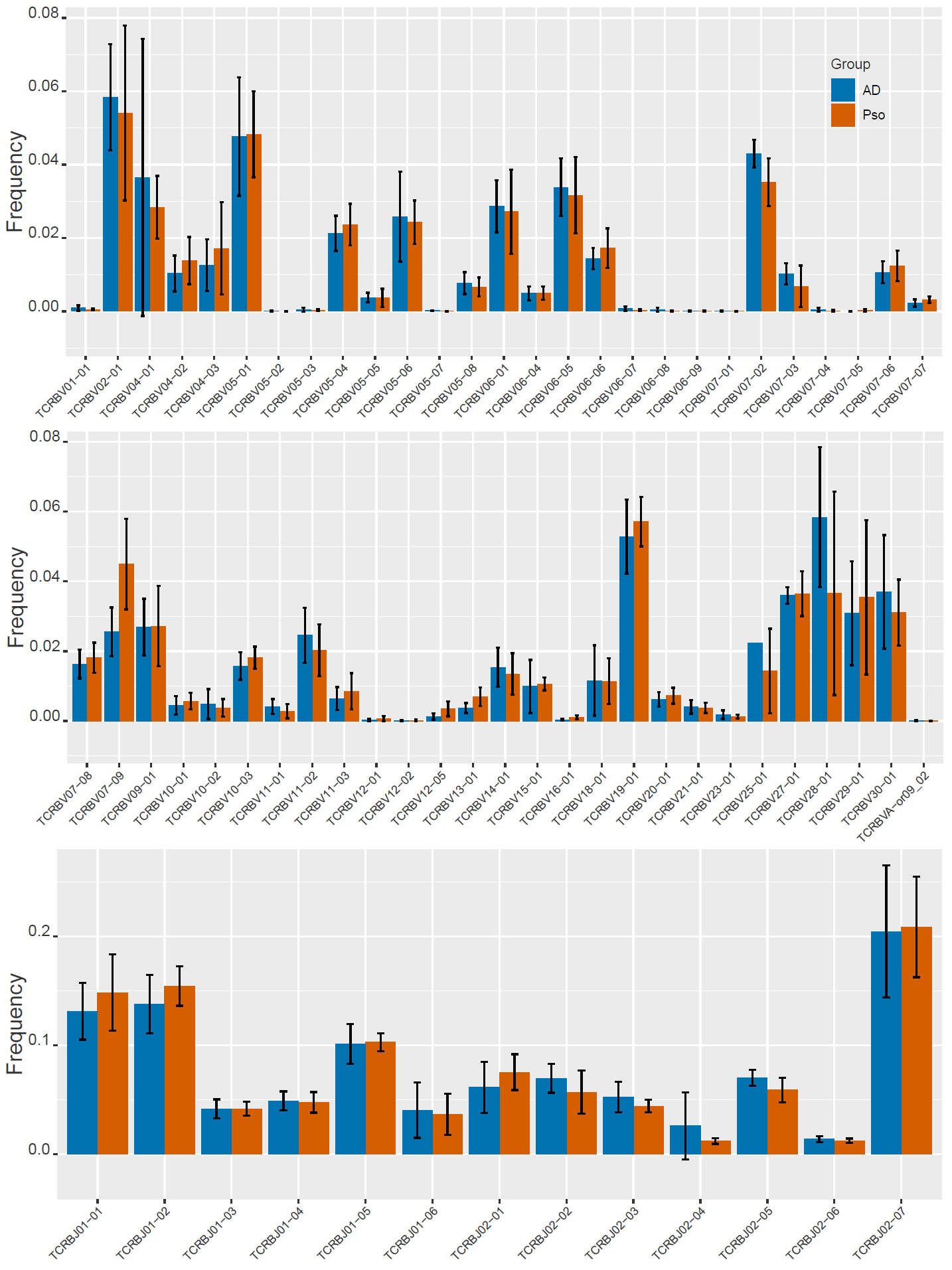

**Supplemental Figure 4**: Percentages of the TRB genes used by skin-homing (CLA^+^) T cells in AD compared to psoriasis. AD n=9, Psoriasis n=10.

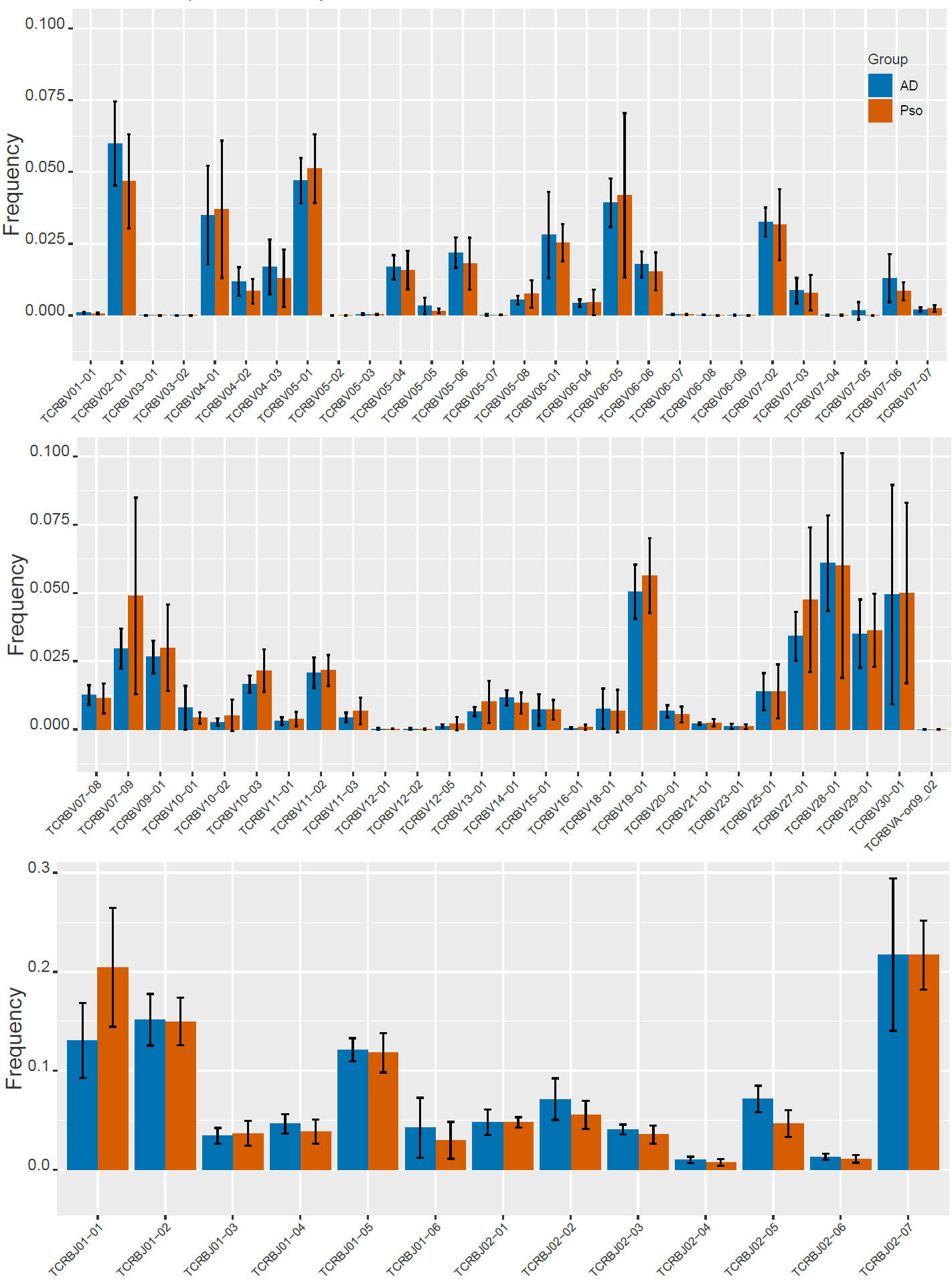
**Supplemental Figure 5**: Percentages of the TRB genes used by non-skin-homing (CLA^-^) T cells in AD compared to psoriasis. AD n=9, Psoriasis n=10.

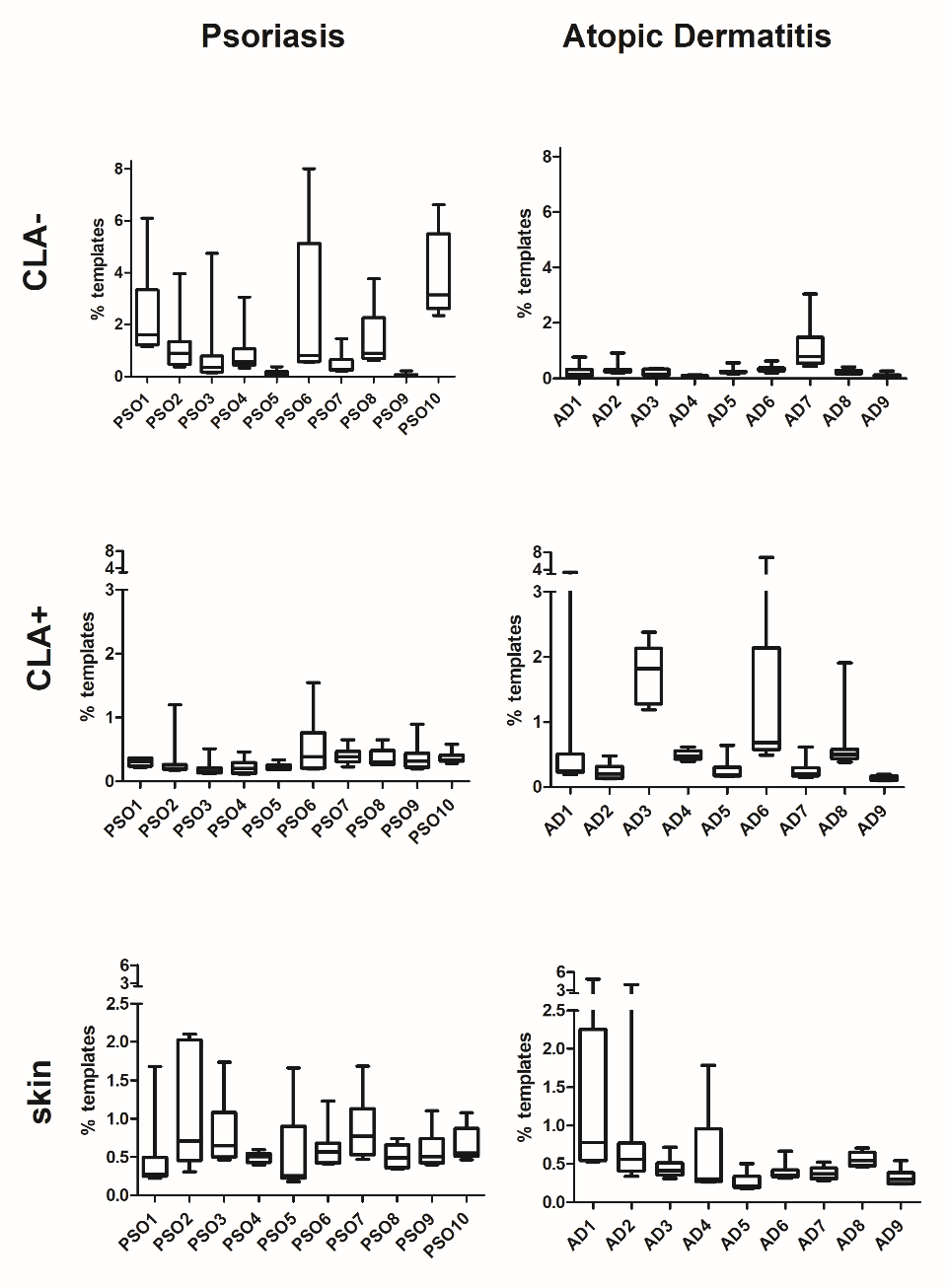

**Supplemental Figure 6**: Combined clonal frequencies of the donors´ 10 most frequent T cell clones. Individual donor data to Figure 3B.

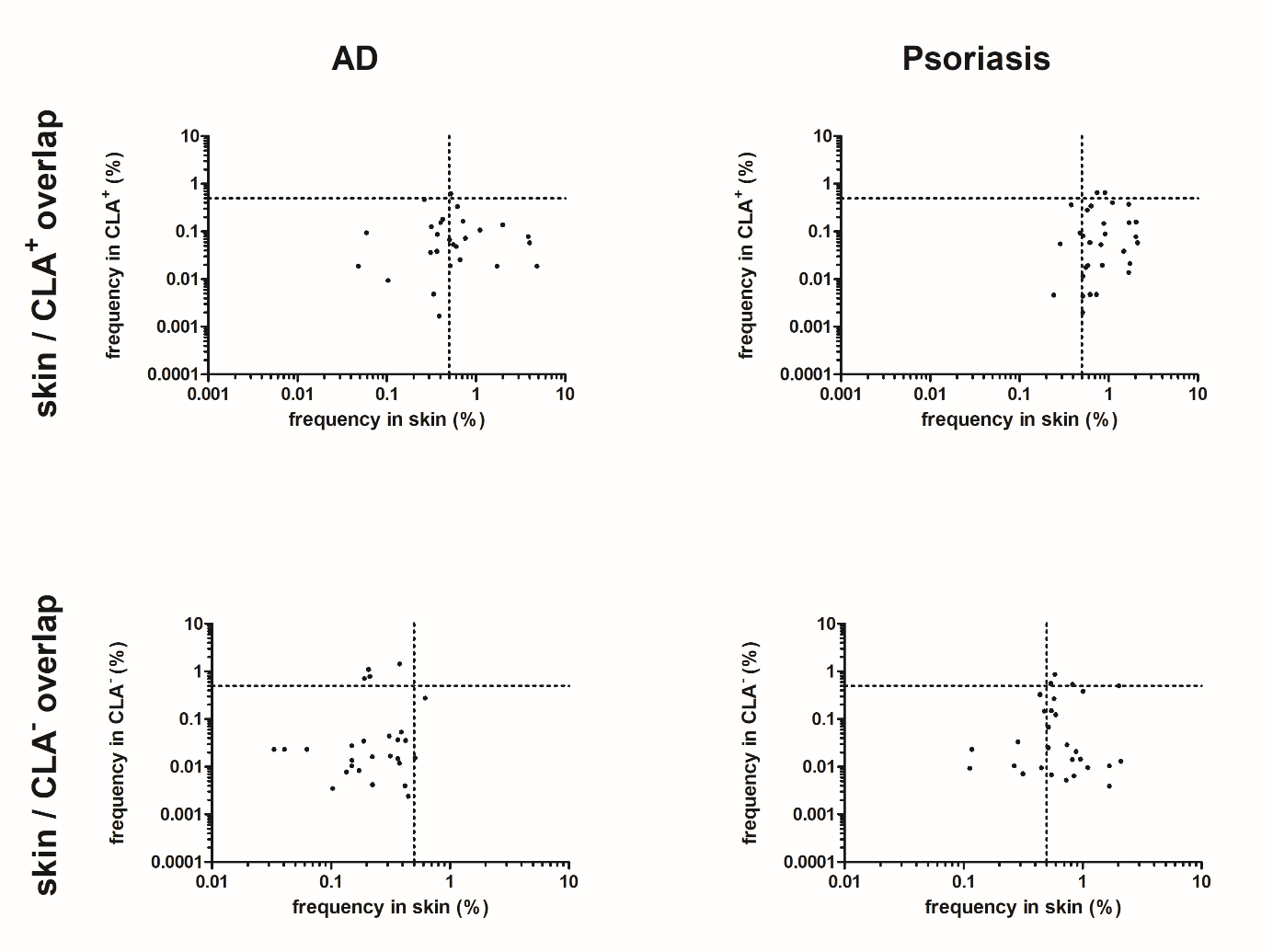

**Supplemental Figure 7:** **Comparative TCR repertoire analysis of highly clonally propagated T cells; frequencies of skin-derived T cell clones that can be detected in either CLA^+^ or CLA^-^ fraction.**

Each plot depicts the frequencies of the three T cell clones with the highest frequency in lesional skin of each donor that are shared between the indicated fractions. The frequencies in the skin are depicted on the x-axis, the frequencies of the respective fraction of circulating cells is depicted on the y-axis.

T cell clones that were detected in both lesional skin and in CLA^+^ fraction are detectable in frequencies above 0.5% in both AD and psoriasis skin. T cell clones that were detected in both lesional skin and in CLA^-^ fraction do not occur in high frequency in AD. In psoriasis, these are often detected in frequencies above 0.5%.

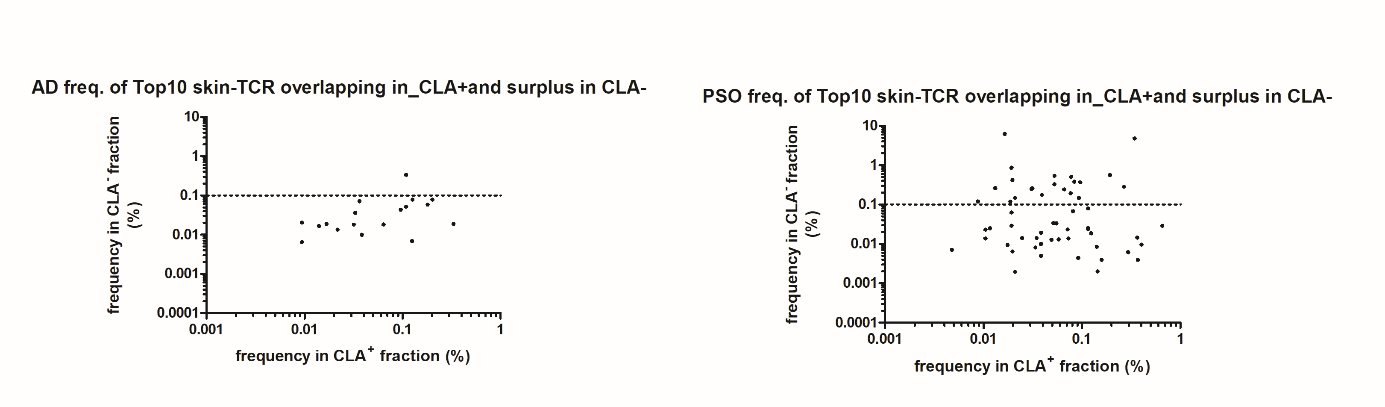

**Supplemental Figure 8. Comparative TCR repertoire analysis of highly clonally propagated T cells; frequencies of skin-derived T cell clones that can be detected in CLA^+^ and surplus CLA^-^ fraction.**

Frequency of the donors´ 10 most frequent T cell clones shared between lesional skin and the CLA^+^ T cell fraction that are also present in the CLA^-^ T cell fraction.

The frequencies within the skin-homing (CLA^+^) fraction of circulating cells are depicted on the x-axis, the frequencies of the skin-homing (CLA^-^) fraction is depicted on the y-axis.

T cell clones that were detected both in lesional skin and in CLA^+^ and surplus in the CLA^-^ fraction are detectable in frequencies above 0.1% in psoriasis but not in AD.
